# Duration of viral shedding and culture positivity with post-vaccination breakthrough delta variant infections

**DOI:** 10.1101/2021.10.14.21264747

**Authors:** Mark J. Siedner, Julie Boucau, Rebecca Gilbert, Rockib Uddin, Jonathan Luu, Sebastien Haneuse, Tammy Vyas, Zahra Reynolds, Surabhi Iyer, Grace Chamberlin, Robert H. Goldstein, Crystal M. North, Chana A. Sacks, James Regan, James P. Flynn, Manish C. Choudhary, Jatin M. Vyas, Amy Barczak, Jacob Lemieux, Jonathan Z. Li

**Author notes:** These authors contributed equally to this manuscript (Amy Barczak, Jacob Lemieux, Jonathan Z. Li).

## Abstract

Isolation guidelines for SARS-CoV-2 are largely derived from data collected prior to emergence of the delta variant. We followed a cohort of ambulatory patients with post-vaccination breakthrough SARS-CoV-2 infections with longitudinal collection of nasal swabs for SARS-CoV-2 viral load quantification, whole genome sequencing, and viral culture. All delta variant infections (8/8, 100%) in our cohort were symptomatic, compared with 64% (9/14) of non-delta variant infections. Delta variant breakthrough infections were characterized by higher initial viral load, longer duration of virologic shedding by PCR (median 13.5 vs 4 days, hazard ratio [HR] 0.45, 95%CI 0.17-1.17), greater likelihood of replication-competent virus at early stages of infection (6/8 [75%] vs 3/14 [23%], *P*=0.03), and longer duration of culturable virus (median 7 vs 3 days, HR 0.38, 95%CI 0.14-1.02) compared to non-delta variants. Nonetheless, no individuals with delta variant infections had replication-competent virus by day 10 after symptom onset or 24 hours after resolution of symptoms. These data support current US Center for Disease Control isolation guidelines and reinforce the importance of prompt testing and isolation among symptomatic individuals with delta variant breakthrough infections. Additional data are needed to evaluate these relationships among asymptomatic and more severe delta variant breakthrough infections.

## Background

Isolation and distancing practices are fundamental elements of COVID-19 epidemic control. Current guidelines authored by the US Centers for Disease Control and Prevention (CDC) recommend most individuals infected with SARS-CoV-2 virus remain isolated for 10 days after a positive test (if asymptomatic) or 10 days from onset of symptoms and 1 day after resolution of symptoms (for symptomatic infections) (1). These guidelines were largely developed based on the low likelihood of recovering replication-competent virus after 10 days of symptoms for most patients (2–5), prior to the emergence of the delta variant as the dominant circulating strain globally (6). The delta variant has been associated with a higher basic reproductive number (7), higher viral loads at detection of infection (8), higher replication efficiency (9), and shorter incubation period and generation time (10). However, there are few longitudinal data on delta variant infections that describe the duration of contagiousness or isolation of replication-competent virus, particularly with post-vaccination breakthrough infections.

## Results and Discussion

Twenty-two individuals with PCR-confirmed SARS-CoV-2 infection after vaccination were enrolled between January and August 2021 (Table 1). All 8 infections (36%) after 29^th^ June 2021 were confirmed by sequencing as delta variant infections. The 14 participants enrolled prior to that date had a diversity of variants including alpha (n=2), gamma (n=1), and mu (n=1). Eight specimens could not be sequenced due to insufficient viral material. Participants with delta and non-delta infections were similar in terms of age, sex, and COVID-19 vaccine manufacturer. Those with delta variant infections had a longer duration of time since completion of vaccination (median 160 vs 29 days), were somewhat more likely to be symptomatic during their course of infection (100 vs 64%) and had a higher viral load at the first study specimen collection (5.5 vs 2.0 log_10_ copies/mL, *P*=0.005).

**Table 1.** Cohort characteristics

|  | Delta Variant<br>Infections<br>(n=8) | Non/Pre-Delta<br>Variant Infections<br>(n=14)<sup>#</sup> | P-value<sup>†</sup> |
| --- | --- | --- | --- |
| Age (mean, SD) | 47 (16) | 41 (16) | 0.41 |
| Female Sex (n %) | 4 (50%) | 8 (57%) | >0.99 |
| Days since full Vaccination* (median, IQR) | 160 (152-185) | 29 (7-54) | 0.002 |
| Vaccine received (n, %) |  |  | 0.27 |
| Moderna | 3 (38%) | 9 (64%) |  |
| Pfizer/BioNTech | 4 (50%) | 5 (36%) |  |
| Johnson & Johnson/Janssen | 1 (13%) | 0 (0%) |  |
| Isolation of replication-competent virus (n, %) | 6 (75%) | 3/14 (21%) | 0.026 |
| Symptomatic infection (n, %) | 8 (100%) | 9 (64%) | 0.115 |
| Culturable virus among those with<br>symptomatic infection (n, %) | 6 (75%) | 3/9 (33%) | 0.153 |
| First study viral load (log10 copies/mL,<br>median, IQR) | 5.6 (4.7 – 6.2) | ND (ND – 5.1) | 0.021 |

SD: standard deviation; IQR: Interquartile range; ND: Not detected
<sup>#</sup>Eight participants with specimens with insufficient virus for sequencing but collected prior to 18<sup>th</sup> June 2021 (when delta first made up more than 10% of sequenced virus in Massachusetts) are presumed pre-delta
\*Two non-delta variant infections had received one dose at the time of infection and were considered to have 0 days from full vaccination
<sup>†</sup>P-values represent statistical tests comparing delta and non-delta variant breakthrough infection characteristics and were estimated using rank sum non-parametric tests for continuous variables (age, days since vaccination, first study viral load) and exact Fischer tests for categorical variables (sex, vaccine received, symptomatic infection, and culturable virus).

Delta variant post-vaccine breakthrough infections were more likely to grow in culture than those with alternate variants (6/8 [75%] vs 3/14 [23%], *P*=0.03). This pattern was consistent when restricted to symptomatic infections only (6/8 [75%] vs 3/9 [33%], *P*=0.03). Individuals with delta variant infections had slower viral load decay assessed by PCR (median time 13.5 vs 4.5 days, HR 0.45, 95%CI 0.17, 1.17, Figures 1 & 2A). Although delta variant infection was also associated with lower hazard of conversion to negative viral culture (HR 0.38, 95%CI 0.14, 1.02), the difference in median time to negative viral culture was less pronounced than it was for conversion to negative PCR (median time to negative viral load 7 vs 4 days, Figure 2B). Seven of 8 (88%) individuals with delta variant post-vaccine breakthrough infections had a confirmed negative viral culture within 10 days of symptom onset. The remaining participant had culturable virus at day 11 but a negative culture at day 13. That participant remained symptomatic at day 11.

**Figure 1.**
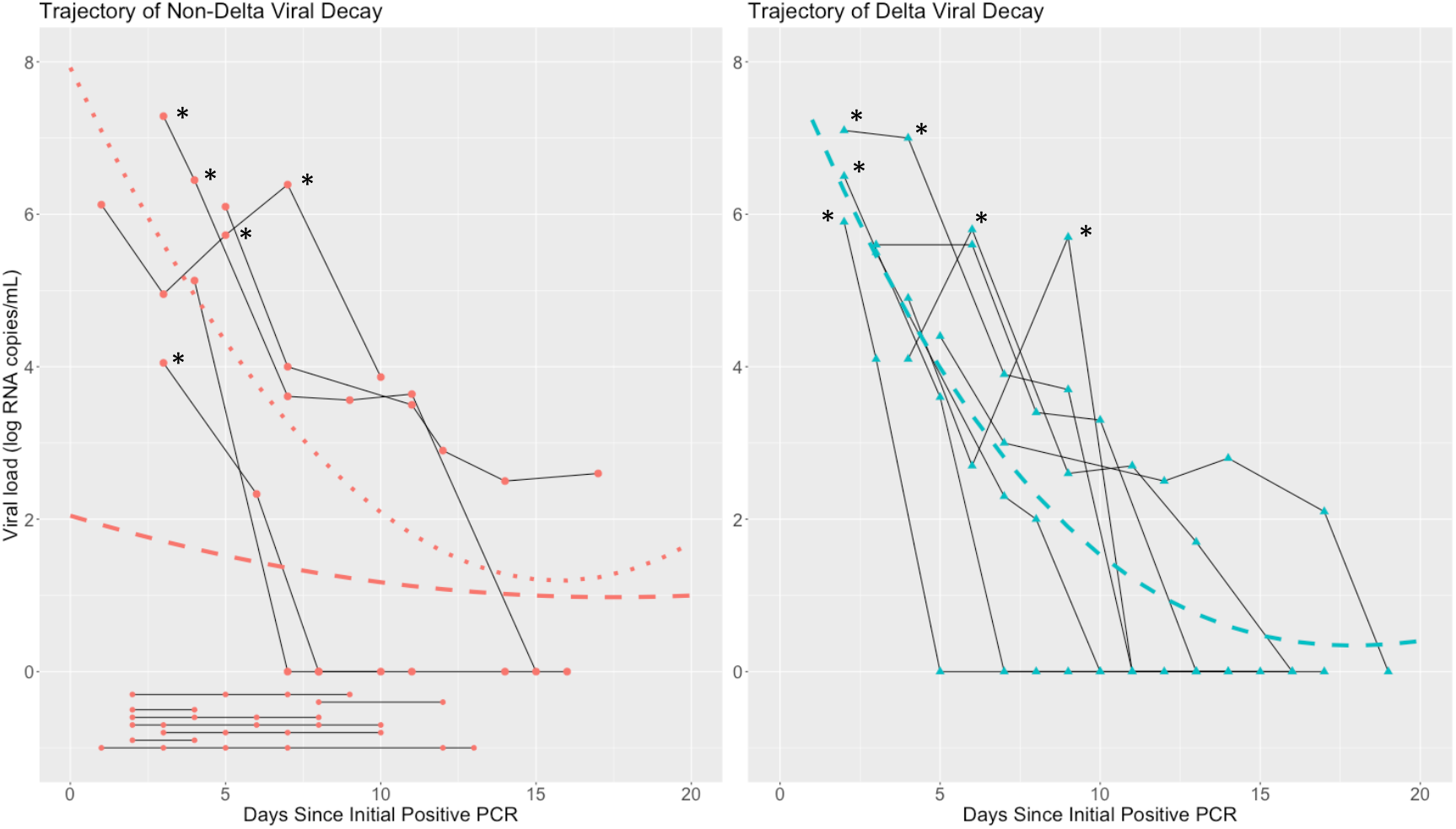
Viral load decay curve in individuals with post-vaccination breakthrough SARS-CoV-2 infection with non-delta (A) and delta variant infections Trajectory of viral load by PCR from time of index positive test for each study participant with non-delta variant (A) and delta variant (B) PCR-confirmed SARS-CoV-2 infection. Each connected solid line represents a participant. The solid dashed lines represent the total population mean line of fit derived from a regression equation including quadratic and cubic terms. The dotted line represents a similar line of fit, but restricted to individuals who remained positive at day 3. Timepoints denoted with asterisks were viral culture positive.

**Figure 2.**
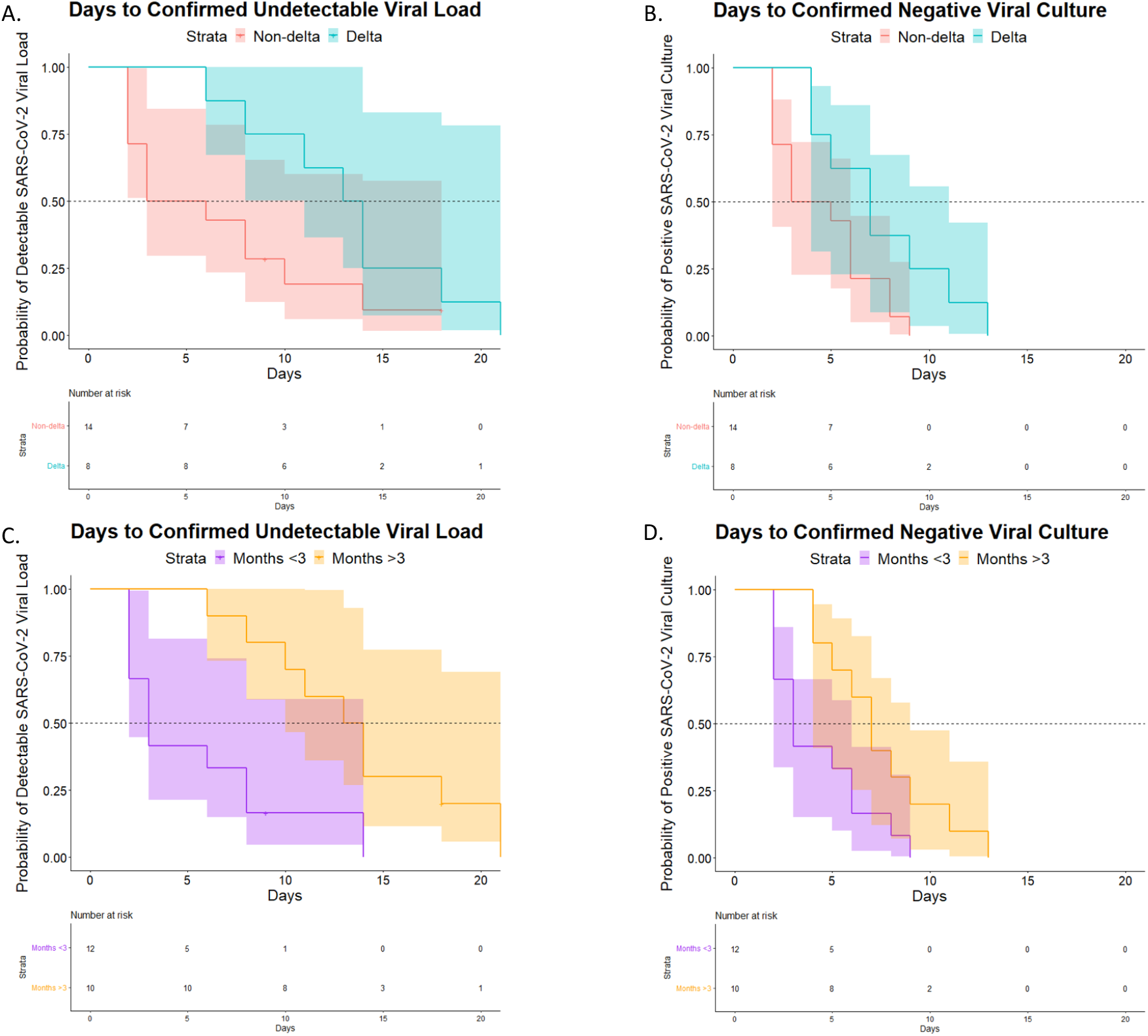
Kaplan-Meier curves indicating days to negative viral load (A) and negative culture (B) by viral variant and days to negative viral load (C) and negative culture (D) by duration of time since completion of COVID-19 vaccination. Observation time begins at the date of positive PCR for asymptomatic cases and date of symptom onset for symptomatic cases.

Similarly, we found evidence that time to negative viral load by PCR was longer for individuals infected more than 3 months after completion of vaccination compared to those infected within 3 months of vaccination (median time 13.5 vs 3 days, HR 0.23, 95%CI 0.08, 0.65, Figure 2C), with significant, albeit diminished, difference in time to negative viral culture (median time 7 vs 3 days, HR 0.37, 95%CI 0.15, 0.92, Figure 2D). When considering time from vaccination as a continuous measure, each additional 30 days since completion of vaccination was associated with an additional 1.3 days of PCR positivity (95%CI -0.2, 2.6 days) and an additional 0.6 days of viral culture positivity (95%CI -0.8, 13 days, Figure 3). Secondary analyses restricted to symptomatic individuals generally showed similar patterns with wider confidence intervals due to the restricted sample size (Supplemental Table and Supplemental Figures 1A-D).

**Figure 3.**
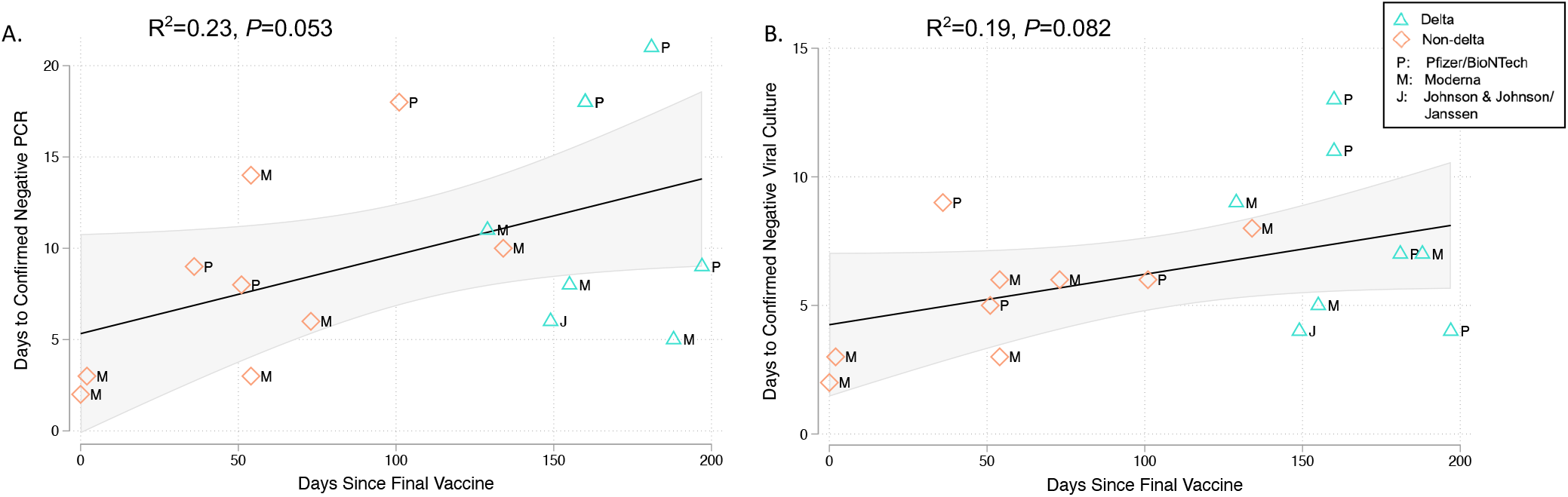
Scatter plots demonstrating relationship between time since completion of COVID-19 vaccination and duration of positive PCR positivity (A) and viral culture positivity (B). Each point on the graph indicates an individual with post-vaccination breakthrough COVID-19 infection. Vaccines received are indicated by plot labels as Moderna (M), Pfizer/BioNTech (P), or Johnson & Johnson/Janssen (J). Black solid lines indicated the line of best fit from linear regression models, whereas gray shaded area indicates the 95% confidence interval around this estimate. R-squared and *P*-values are estimates from these models. Green triangles indicate delta variant infections whereas blue diamonds represent non-delta variant infections.

In this cohort of ambulatory individuals with post-vaccination breakthrough infections, symptomatic delta variant SARS-CoV-2 infections were characterized by high initial viral load and a longer duration of virologic shedding as detected by PCR (median 13.5 days vs 4.5 days). Moreover, identification of replication-competent virus by culture was more common with delta variant infections than non-delta infections (75% vs 21%) and the duration of replication-competent virus was modestly prolonged among those with delta variant breakthrough infections (7 days vs 3 days). Nonetheless, we detected only a single breakthrough infection characterized by more than 10 days of replication-competent virus (11 days) in a participant who remained symptomatic on their final day of culture positivity (day 11).

Unlike our study, a prior study showed similar trajectories and duration of virologic shedding as detected by PCR between delta and alpha viral variants, and shorter duration of shedding among delta variant infections than we found (median 6 vs 13.5 days) (11). We suspect that this difference is explained by distinct features of our study populations. Whereas that former study included relatively young individuals being tested as part of their affiliation with a sports league, study subjects with breakthrough delta infections in our study population were comparatively older adults and accessing SARS-CoV-2 testing through a healthcare system.

Our data are in keeping with current guidelines recommending isolation for 10 days or until symptom resolution for symptomatic post-vaccination breakthrough infections and add new evidence in support of these guidelines in the delta variant era. These results also reinforce that post-vaccination breakthrough delta infections should be considered contagious and highlight the critical importance of prompt testing for symptomatic vaccinated individuals due to the high frequency of identification of replication-competent virus (5, 12).

Our study was limited to a small sample (n=22), to those vaccinated, and to non-severe infections in ambulatory individuals. We did not perform ongoing contact tracing in our study. Therefore, our conclusions about contagiousness are limited to inferences about the presence or absence of replication-competent virus, but not confirmation of downstream infections from the cases detected in our study. All delta variant infections in our cohort were mild, but symptomatic, and thus our results should not be generalized to asymptomatic infections. Because of collinearity between delta variant infections and duration of time between vaccination and infection in our cohort, we cannot meaningfully distinguish the relative contributions of these two factors on transmission dynamics in breakthrough infections. Importantly, additional data are also needed to better elucidate the dynamics of delta variant infection in unvaccinated individuals. Work prior to the emergence of the delta variant suggests that the magnitude of viral load and duration of viral shedding is more prolonged among unvaccinated individuals (11, 13). That data in combination with ours, which demonstrates longer shedding of nucleic acid and isolation of replication-competent virus in symptomatic delta variant-infected individuals, suggests that current isolation guidelines might not be adequate for all unvaccinated individuals with delta variant infections. Additional work is also needed to assess transmission dynamics in individuals with severe infection and after vaccine booster administration.

## Methods

### Study Participants

We enrolled non-hospitalized individuals with confirmed SARS-CoV-2 infection after vaccination. Participants were recruited after positive tests through one of two means. First, between January and March of 2021, employees in the Mass General Brigham Medical System were offered weekly COVID-19 testing irrespective of symptoms (14). Following the conclusion of that program, we began recruiting all individuals with positive SARS-CoV-2 PCR test results in the Mass General Brigham Medical System, which includes testing for symptomatic individuals as well as asymptomatic testing for contact tracing and screening procedures (e.g. pre-operative clearance). All adults over 18 who tested positive by either of these systems were eligible for inclusion in this study. For those who consented to participation, we conducted home visits three times weekly until negative PCR testing. At each visit, we obtained self-collected nasal swabs for SARS-CoV-2 PCR, culture and whole genome sequencing. Symptoms were assessed at each specimen collection and through medical chart review after study completion. Symptomatic infections were defined as those with COVID-19-related symptoms at any point during the observation period.

### Viral Load Quantification

Viral load quantification and sequencing was conducted as previously reported (14). Briefly, we pelleted virions from nasal swab fluids after centrifugation at 21,000 x g for 2 hours at 4°C. We added TRIzol-LS™ Reagent (ThermoFisher) to the pellets and incubated the pellets on ice after removing the supernatant. We then vortexed the pellets in 200 μL of chloroform (MilliporeSigma), centrifuged the mixtures at 21,000 x g for 15 minutes at 4°C, removed the aqueous layer, and then treated the resulting solution with an equal volume of isopropanol (Sigma). We then added GlycoBlue™ Coprecipitant (ThermoFisher) and 100 μL 3M Sodium Acetate (Life Technologies) and incubated the mixtures in dry ice. We produced RNA pellets by centrifugation at 21,000 x g for 45 minutes at 4°C, discarded the supernatant, washed the RNA with cold 70% ethanol, and resuspended it in DEPC-treated water (ThermoFisher). We quantified SARS-CoV-2 RNA virus using RT-qPCR with the US CDC 2019-nCoV_N1 primer and probe set (IDT) (15). Reactions included extracted RNA, 1X TaqPath™ 1-Step RT-qPCR Master Mix, CG (ThermoFisher), forward and reverse primers, and the probe. We quantified viral copy numbers using N1 qPCR standards in 16-fold dilutions to generate standard curves. Each sample was run in triplicate with two non-template control (NTC) wells that were included as negative controls. Additionally, we tested positive and negative controls alongside all samples. We assessed sample quality by quantifying the Importin-8 (IPO8) housekeeping gene RNA level. Finally, to determine the efficiency of RNA extraction and qPCR amplification, we spiked (RCAS)(16) into each sample as an internal virion control.

### SARS-CoV-2 Whole Genome Sequencing

We performed whole genome sequencing using the Illumina COVIDSeq Test protocol. We constructed libraries using the Illumina Nextera XT Library Prep Kit, then pooled and quantified the libraries using a Qubit High Sensitivity dsDNA kit. Then, we performed genomic sequencing on an Illumina NextSeq 2000, Illumina NextSeq 550, or Illumina NovaSeq SP instrument. Sequences with an assembly length greater than 24000 base pairs were considered complete genomes, and we assigned those sequences a Pango lineage using the most up-to-date version of pangoLEARN assignment algorithm.

### SARS-CoV-2 Spike gene amplification

We additionally performed spike gene amplification, as previously described (14), to determine variant types for specimens with low viral load to determine variants when whole genome sequencing was unsuccessful. We used Superscript IV reverse transcriptase (Invitrogen, Waltham, MA, USA) to conduct cDNA synthesis. To exclude PCR artifacts, we used two strategies to amplify the SARS-CoV-2 spike gene: 1) Nested PCR amplification with *in-house* designed primer sets that targeted codon 1-814 of the spike gene and 2) the multiplexed primer pools designed with Primal Scheme generating 400-bp tiling amplicons based on the Arctic protocol (17). We separately pooled PCR products from both strategies and performed Illumina library construction using the Nextera XT Library Prep Kit (Illumina, San Diego, CA, USA). We analyzed raw sequence data with PASeq v1.4 (https://www.paseq.org). We conducted data filtering with Trimmomatic (v0.30) (18), using min a Q25/5 bp sliding window and a 70 bp minimum length. We filtered out non-viral contamination with BBsplit v35.76 (19). We then merged filtered reads using paired-end read merger v0.9.6 aligned to reference sequences with Bowtie2 v2.1.0) (20). Finally, amino acid variants were identified at the codon level with perl code and used to determine SARS-CoV-2 variant type.

### SARS-CoV-2 culture

We performed viral culture as previously reported in the BSL3 laboratory of the Ragon Institute of MGH, MIT, and Harvard (14, 21). Briefly, we detached Vero-E6 cells (ATCC) maintained in DMEM (Corning) supplemented with HEPES (Corning), 1X Penicillin/Streptomycin (Corning), 1X Glutamine (Glutamax, ThermoFisher Scientific), and 10% Fetal Bovine serum (FBS) (Sigma) using Trypsin-EDTA (Fisher Scientific) and seeded the cells at 75,000 cells per wells in 24w plates or 20,000 in 96w plates 16-20 hours before infection. We thawed specimens on ice, filtered the specimens through a Spin-X 0.45um filter (Corning) at 10,000 x g for 5min, and diluted them 1:10 in DMEM supplemented with HEPES, 1X Penicillin/Streptomycin and 1X Glutamine. We used 100uL of the solution to inoculate triplicate wells in a 24 well plate. We then added 1mL of DMEM supplemented with HEPES, 1X Penicillin/Streptomycin and 1X Glutamine and 2% FBS to each well after 1h of incubation and removal of the viral inoculum. We added 25ul of the undiluted filtrate to four wells of a 96w plate and serial diluted (1:5) the filtrate in media containing 5ug/mL of polybrene (Santa Cruz Biotechnology). We centrifuged the 96w plates for 1 hour at 2000 x g at 37C. As a positive control, we used the SARS-CoV-2 isolate USA-WA1/2020 strain (BEI Resources). We observed viral culture plates at 3- and 7-days post-infection with a light microscope and documented wells showing CPE. Lastly, we harvested the supernatant of wells displaying CPE 10-14 days post-infection and isolated RNA using a QIAamp Viral RNA Mini kit (QIAGEN) for confirmation of the viral sequence.

### Statistical Methods

We first evaluated patient-specific trajectories of quantitative viral load by PCR over time since index positive test by constructing spaghetti plots. To estimate grouped mean trajectories, stratified by variant, we fitted a linear regression model with quadratic and cubic spline terms. We then used the Kaplan-Meier estimator to estimate the survivor function for: (1) time to negative viral by PCR testing and (2) time to negative viral culture. For both outcomes, we took the time of symptom onset (for symptomatic cases) or time of first positive PCR (for asymptomatic cases) as the origin of the timescale. We selected these definitions of left censoring to replicate US CDC isolation guidelines. For both outcomes, we estimated survivor functions stratified by delta versus non-delta variant infection and, separately, by duration of time from vaccination to infection, dichotomized as greater versus less than 90 days. We next fitted cox proportional hazards model with both outcomes and delta versus non-delta variant infection and, separately, time since completion of vaccination as predictors. In sensitivity analyses to assess for potential confounding by the presence of absence of symptoms, we re-estimated Kaplan-Meier survivor functions after limiting the sample to individuals with symptomatic infection. Finally, we fit linear regression models and plotted scatter plots to assess relationships between duration of PCR and culture positivity and time since completion of vaccination as a continuous measure.

### Study Approval

Study procedures were reviewed and approved by the Human Subjects Institutional Review Board and the Institutional Biosafety Committee at Mass General Brigham. All participants gave verbal informed consent, as written consent was waived by the review committee based on the risk to benefit ratio of requiring in-person interactions for an observational study of COVID-19.

## Data Availability

Deidentified data are available upon request to the corresponding author.

## Author contributions

MJS conceived of the study design, participated in the data analysis, drafted the initial manuscript and contributed editorial input.

JB conducted experiments, participated in the data analysis, and contributed editorial input.

RG directed the data collection, participated in the data analysis, and contributed editorial input.

RU participated in data collection, data analysis, and contributed editorial input

JL participated in the data analysis and contributed editorial input. SH participated in the data analysis and contributed editorial input.

TV participated in the data collection and contributed editorial input.

ZR participated in the data collection and contributed editorial input.

SI participated in data collection and contributed editorial input.

GC participated in data collection and contributed editorial input.

RG participated in data collection and contributed editorial input.

CMN participated in data collection and contributed editorial input.

CAS participated in data collection and contributed editorial input.

JR participated in the data collection, conducted experiments and contributed editorial input. JF conducted experiments and contributed editorial input.

MC conducted experiments, participated in the data analysis and contributed editorial input.

JV conceived of the study design, provided reagents and contributed editorial input.

AB conceived of the study design, provided reagents, participated in the data analysis, and contributed editorial input.

JL conceived of the study design, provided reagents, participated in the data analysis, and contributed editorial input.

JZL conceived of the study design, provided reagents, participated in the data analysis, and contributed editorial input.

All authors approve of the final version of the manuscript.

## Acknowledgments

The study team would like to thank Dr. Marcia Goldberg for her contributions to this work.

