## Supplemental Table 1 and Supplemental Figure 1 for "Duration of viral shedding and culture positivity with post-vaccination breakthrough delta variant infections"

**Supplementary Material.**

**Supplement Table 1.** Median time and hazard of negative viral load by polymerase chain reaction and negative viral culture by variant and symptomatic infection……………………………….2

**Supplement Figure 1.** Kaplan-Meier curves, restricted to symptomatic cases only, indicating days to negative viral load (A) and negative culture (B) by viral variant and days to negative viral load (C) and negative culture (D) by duration of time since completion of COVID-19 vaccination. Observation time begins on the date of symptom onset……………………………………….3

**Supplemental Table 1**. Median time and hazard of negative viral load by polymerase chain reaction and negative viral culture by variant and symptomatic infection.

| | **Delta Variant Infections (n=8)** | **All Non-Delta Variant Infections (n=14)** | **Hazard Ratio (95%CI) Delta vs all Non-Delta Infections** | **Symptomatic Non-Delta Infections (n=9)** | **Hazard Ratio (95%CI) Delta vs Symptomatic Non-Delta Infections** |
|---|---|---|---|---|---|
| Median time to negative viral load by PCR (days) | 13.5 | 4.5 | 0.45 (0.17, 1.17) | 8 | 0.62 (0.21, 1.78) |
| Median time to negative viral culture (days) | 7 | 4 | 0.38 (0.14, 1.02) | 6 | 0.44 (0.15, 1.28) |
| | **>3 months since vaccination (n=10)** | **All <3 months since vaccination (n=12)** | **Harvard Ratio (95%CI) >3 months vs All <3 months** | **Symptomatic and <3 months since vaccination (n=7)** | **Harvard Ratio (95%CI) >3 months vs Symptomatic <3 months** |
| Median time to negative viral load by PCR (days) | 13.5 | 3 | 0.23 (0.08, 0.65) | 6 | 0.31 (0.10, 0.97) |
| Median time to negative viral culture (days) | 7 | 3 | 0.37 (0.15, 0.92) | 5 | 0.41 (0.15, 1.16) |

**Supplemental Figure 1.** Kaplan-Meier curves, restricted to symptomatic cases only, indicating days to negative viral load (A) and negative culture (B) by viral variant and days to negative viral load (C) and negative culture (D) by duration of time since completion of COVID-19 vaccination. Observation time begins on the date of symptom onset

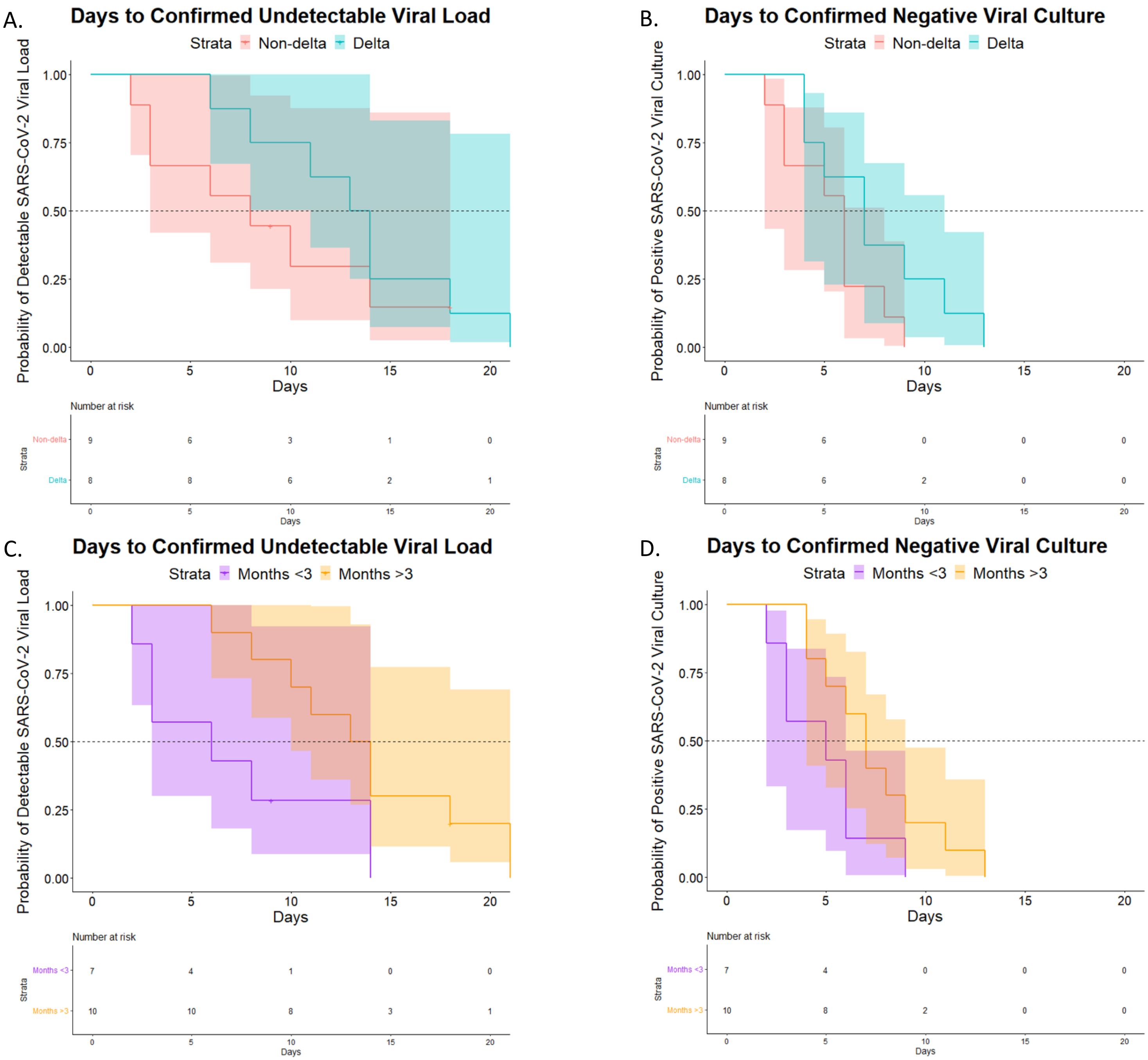
